# A conceptual model for pluralistic healthcare behavior: results from a qualitative study in southwestern Uganda

**DOI:** 10.1101/19008508

**Authors:** Radhika Sundararajan, Juliet Mwanga-Amumpaire, Rachel King, Norma C. Ware

**Affiliations:** Department of Emergency Medicine, Weill Cornell Medicine, 525 East 68^th^ Street, New York, 10065, USA; Weill Cornell Center for Global Health, New York, USA; Mbarara University of Science and Technology, Mbarara, Uganda; Global Health Sciences, University of California San Francisco, USA; Department of Medicine, Brigham and Women’s Hospital, Boston, USA; Department of Global Health and Social Medicine, Harvard Medical School, Boston USA

**Keywords:** Medical pluralism, Uganda, traditional healers, qualitative

## Abstract

**Introduction:** *Medical pluralism*, or concurrent utilization of multiple therapeutic modalities, is common in various international contexts, and has been characterized as a factor contributing to poor health outcomes in low-resource settings. Traditional healers are ubiquitous providers in most regions, including the study site of southwestern Uganda. It is not well understood why patients in pluralistic settings continue to engage with *both* therapeutic healthcare modalities, rather than simply selecting one or the other. The goal of this study was to identify factors that motivate pluralistic healthcare utilization, and create a general, conceptual framework of pluralistic health behavior.

**Methods:** In-depth interviews were conducted between September 2017 and February 2018 with patients seeking care at traditional healers (N=30) and at an outpatient medicine clinic (N=30) in Mbarara, Uganda; the study is nested within a longitudinal project examining HIV testing engagement among traditional healer-utilizing communities. Inclusion criteria included age ≥18 years, and ability to provide informed consent. Participants were recruited from healer practices representing the range of healer specialties. Following an inductive approach, interview transcripts were reviewed and coded to identify conceptual categories explaining healthcare utilization.

**Results:** We identified three broad categories relevant to healthcare utilization among study participants: 1) traditional healers treat patients with “care”; 2) biomedicine uses “modern” technologies; and 3) peer “testimony” influences healthcare engagement. These categories describe variables at the <u>healthcare provider</u>, <u>healthcare system</u>, and <u>peer</u> levels that interrelate to motivate individual engagement in pluralistic health resources.

**Conclusions:** Patients perceive clear advantages and disadvantages to biomedical and traditional care in medically pluralistic settings. We identified factors at the healthcare provider, healthcare system, and peer levels which influence patients’ therapeutic itineraries. Our findings provide a basis to improve health outcomes in medically pluralistic settings, and underscore the importance of recognizing traditional healers as important stakeholders in community health.

**STRENGTHS AND LIMITATIONS OF THIS STUDY**

- Medical pluralism is common in both high- and low-resource settings, and has been characterized as a factor leading to poor health outcomes for both infectious and non-communicable diseases
- This study identifies factors that motivate utilization of healthcare in a medically pluralistic community
- Patients in pluralistic settings perceive clear advantages and disadvantages of both traditional care and biomedicine; characteristics of healthcare providers, the healthcare system, and peer influences motivate patients to engage with particular healthcare modalities
- Patients often prefer traditional healing instead of biomedicine; this utilization is not simply a function of limited access to biomedical resources
- Traditional healers should be considered important stakeholders in community health

## INTRODUCTION

*Medical pluralism*, or utilization of multiple therapeutic modalities, is common where both biomedical and complementary or alternative treatments are available to patients. This pattern of healthcare engagement is observed in both high-[1,2] and low-resource settings[3-5], and is well described for patients with both acute[6-8] and chronic illness[3,9-11] in various international contexts.

In low- and middle-income countries, traditional medicine is utilized instead of, or in concert with, biomedical therapies. Traditional healers have been defined by the World Health Organization as: 1) persons recognized by local community as healers; 2) having regular patient attendance; and 3) having space to receive and treat patients[5]. They “provide health care by using plant, animal and mineral substances, and other methods based on social, cultural, and religious practices” [12]. Prior work in medically pluralistic contexts shows that initial choice of therapeutic modality is driven by patients’ perceived etiology of illness, and provider trustworthiness[13-16]. Patients may switch modalities in the setting of treatment “failure”, when symptoms worsen or persist despite treatment[13,17].

Medical pluralism has been characterized as a central factor contributing to poor health outcomes. For example, researchers have shown that use of traditional medicine delays HIV testing and ART initiation[18], and interrupts HIV treatment[13], for people living with HIV (PLHIV). In Mozambique, PLHIV initially seeking care from traditional healers experienced significantly longer delays to diagnosis compared with those who did not utilize healers; this delay exponentially increased with corresponding increases in the number of healers consulted prior to receiving HIV testing[18]. In South Africa, medical pluralism was shown to be negatively associated with ART use in a cohort of PLHIV[19]. Research has also demonstrated that medical pluralism contributes to poor outcomes for non-infectious diseases, such as nonadherence to chemotherapy for cancer^4,11^, or poor outpatient linkage to care for patients with hypertension[11].

In many parts of the world, traditional healers are extensively utilized; for example, it is estimated that 80% of the population in sub-Saharan Africa visit traditional healers[5]. Traditional healer utilization may be partially attributed to accessibility: healers are present in higher numbers than physicians and biomedical resources, particularly in low- resource settings[5]. However, their popularity cannot be strictly explained by convenience; research in urban regions having high density of biomedical facilities demonstrates similar reliance on traditional healers[1,3]. Patients also seek traditional therapies to address symptoms attributed to ancestral curses or bewitching, believed incurable by biomedicine[20]. Use of traditional medicine is also strongly tied to local religious and ethnic identities[21].

Biomedical and traditional healers offer distinctive forms of healthcare for patients. In medically pluralistic contexts, it is not well understood why patients continue to engage with *both* therapeutic healthcare modalities, rather than simply selecting one or the other. There have been many disease-specific studies that describe factors influencing pluralistic therapeutic itineraries[17,19,22], but there remains a dearth of knowledge on variables that shape healthcare engagement generally in these communities. The goal of this study was to identify factors that motivate pluralistic engagement with healthcare resources, using qualitative research methods. We sought to characterize salient perceived advantages and disadvantages of each modality, and explain pluralistic therapeutic itineraries in a sub-Saharan African context. These data were used to develop a general, conceptual framework that can inform future research on pluralistic health behavior.

## METHODS

### Study Setting and Design

This qualitative study was conducted in Mbarara District, Uganda, a district of 418,000 residents located ∼275 km southwest of the capital city of Kampala. Southwestern Uganda is a medically pluralistic context, where both traditional and biomedical modalities of healthcare co-exist [23-25]. In this region of sub-Saharan Africa, traditional healers practice herbalism and spiritual healing; they also set broken bones and attend births in the community. Spiritual healers attribute their powers to the *Bachwezi*, which are believed to be ancestral spirits from an ancient kingdom that previously occupied this region of eastern Africa[26,27]. This qualitative study was conducted as part of a multi-year, mixed methods study of HIV services engagement in a medically pluralistic community.

### Sampling and Recruitment

Following a purposive sampling strategy, sixty (N=60) adults were identified to participate as key informants in this study, or “individuals that are especially knowledgeable about or experienced with a phenomenon of interest” [28]. In our case, key informants were selected to represent variation in experiences of receiving modalities of healthcare: biomedical and traditional. That is, participants were patients representing two subgroups: (1) individuals receiving treatment from traditional healers (N=30), and (2) individuals receiving treatment from a biomedical general medicine outpatient clinic (N=30). Inclusion criteria for all participants were: 1) age ≥18 years; 2) ability to provide informed consent; and 3) seeking healthcare at either a traditional healer or outpatient biomedical clinic in Mbarara District.

A target sample size of thirty participants per subgroup was guided by prior research suggesting that a range between 20 and 30 interviews is adequate to reach *thematic saturation*, the point at which no new concepts emerge from subsequent interviews[29- 31]. Two authors (RS and JMA) reviewed transcripts as they were completed and corresponded weekly to identify and discuss emerging themes. After twenty-five interviews per group were conducted, the two authors agreed that interview content no longer contained new or surprising content. Five additional interviews per group were conducted to confirm thematic saturation.

Participants in the traditional medicine subgroup were recruited from twelve traditional healer practices which reflected the range of specialties in this region (herbalist, bone setter, traditional birth attendant and spiritual healer). For the purposes of this study, we excluded Christian-based spiritual healers (i.e., “Born Again” or Pentecostal ministers). Participants in the biomedical subgroup were recruited from Mbarara Municipality Clinic, a general outpatient government-run clinic in the city of Mbarara, which serves approximately 50,000 patients per year.

Healers gave permission for study staff to recruit patients at their practices. At both traditional and biomedical facilities, research assistants approached patients following completion of visits healing sessions to assess eligibility and interest in participation. Recruitment was carried out over a period of six months (September 2017 - February 2018);

### Data Collection

Data collection for this study consisted of a single in-depth interview, conducted by Ugandan research assistants (RAs) trained in qualitative research methods. Interviews followed an interview guide that included the following topics: 1) details of the patient’s therapeutic itinerary for his/her current symptoms; 2) symptoms that motivated him/her to seek healthcare; 3) attitudes towards, and experiences with, traditional and biomedicine; and 4) details of concurrent or recent biomedical and traditional healer visits. Interviews lasted approximately one hour and were conducted in the local language (Runyankore), in private locations at either healer practices or at the participating biomedical clinic. Participants received the equivalent of 10,000 Ugandan Shillings (UGX, ∼$3 USD) in household staples (cooking oil, sugar, salt, soap) in recognition the time and effort required to participate in the interview.

Interviews were digitally recorded, then transcribed and translated into English by the same RA who had conducted the interview. All transcripts were produced within 72 hours of the interview being completed. The transcripts were reviewed by the first author for quality, content, and to provide feedback to the RAs regarding interviewing techniques. English transcripts were spot-checked against audio recordings by an author (JMA, who is fluent in Runyakore and English) to ensure validity and integrity of translations.

### Analysis of Data

A three-step, inductive approach was used to analyze the qualitative data, as follows: (1) development of codes; (2) coding; and (3) category construction.

#### Development of Codes

Following an inductive approach to qualitative data analysis, interview transcripts were reviewed by the first author (RS) concurrently with data collection to identify an initial set of codes, or labels that described key concepts in the dataset. The inductive strategy provided overlap between qualitative interviewing and data analysis, allowing for iterative engagement with the dataset to identify emerging concepts of interest. As additional transcripts were produced and reviewed, codes were reviewed and refined to fit the data. Using the “constant comparison” method, newly coded text segments were compared to text segments previously marked with the same code to determine if they reflected the same concept[32]. This process was repeated until all transcripts had been reviewed. A final list of codes was produced through discussion and consensus among three co-authors (RS, JMA and RK).

#### Coding

All study transcripts were coded, and re-coded when necessary, using the finalized list of codes. QSR NViVo 11 (QRS International Pty Ltd) was used for coding and data organization, but not in development of codes.

#### Category Construction

Next, coded data were examined and grouped to form conceptual categories, where data are aggregated based on similarities of meaning. Categories are defined below using text examples. Quotes from participants are shown as italicized and indented. Interrelationships between categories were identified to create a conceptual framework illustrating factors that influence pluralistic health behavior (Figure 1).

**Figure 1.**
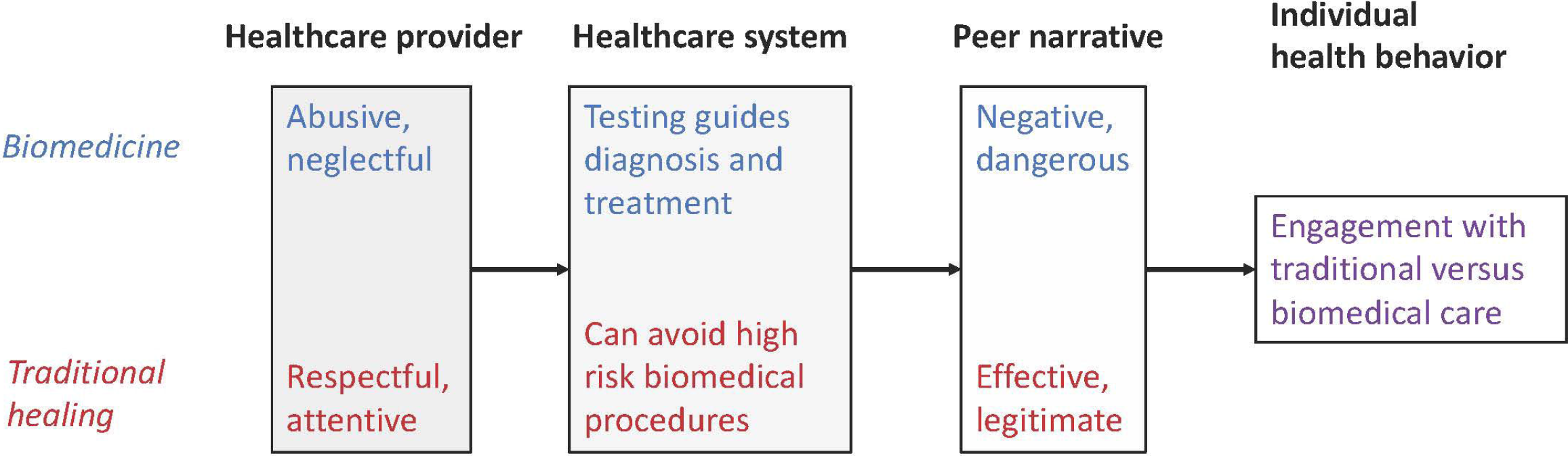
Conceptual model showing key factors within various levels (healthcare provider, healthcare system, peer) influencing individual health behavior within medically pluralistic contexts.

### Ethical Statement

This research was approved by the Human Research Protections Program Institutional Research Board at the University of California, San Diego (#170672), Weill Cornell Medical College (#1803019105), Mbarara University of Science and Technology Research Ethics Committee (#16/01-17) and the Ugandan National Council for Science and Technology (#SS4338). Participants provided written and verbal informed consent in Runyankore.

## RESULTS

### Characteristics of Participants

Characteristics of study participants appear in Table 1. Over half of the sample had clinical experience with both biomedical and traditional modalities of healthcare. However, pluralistic behaviors were much more common among patients of traditional healers. The vast majority of participants recruited from the biomedical clinic denied prior experience receiving care from traditional healers (n=28/30, 93%); in contrast, *all* (n=30) traditional healer patients report prior experience receiving biomedical treatment.

**Table 1.** CHARACTERISTICS OF STUDY PARTICIPANTS.

| TABLE 1. CHARACTERISTICS OF STUDY PARTICIPANTS |  |  |
| --- | --- | --- |
| Characteristic | Traditional healer clients (N=30) | Biomedical clients (N=30) |
| Had previously received care from alternate modality | N=30 (100%) | N=2 (7%) |
| Age (in years) | 36.7 (mean) | 31.6 (mean) |
| Female gender (%) | N = 16 (53%) | N= 18 (60%) |
| Primary school education or less | N= 14 (47%) | N = 13 (43%) |
| Household size (in persons) | 5.4 (mean) | 5.3 (mean) |
| Marital status | Single (N = 7)<br>Married/Cohabiting (N = 21)<br>Widowed (N = 2) | Single (N = 11)<br>Married/Cohabiting (N = 17)<br>Widowed (N = 2) |
| Christian religion | N = 25 | N = 23 |
| Monthly household income (in USD) | \$121 (mean) | \$45 (mean) |
| Type of healer visited | Spiritualist (N=12)<br>Bone setter (N=10)<br>Traditional birth attendant (N=4)<br>Herbalist (N=4) | N/A |

Participants recruited from healer practice locations were slightly older, with a higher proportion being married, and with higher reported monthly incomes, compared to the biomedicine group. Other characteristics, including gender, household size, highest level of education, and religious affiliation, were similar between the two groups.

### Qualitative Results

#### Overview

Our qualitative data demonstrate salient perceived advantages and disadvantages to both healthcare modalities, which motivate patient engagement with healthcare resources. We have developed three broad categories representing influences on healthcare utilization that were evident in the data. They are summarized as follows: 1) traditional healers treat patients with “care”; 2) biomedicine uses “modern” technologies; and 3) peer “testimony” influences healthcare engagement. Within each of these categories, we provide examples to illustrate how these factors drive plural healthcare engagement. We consider each one separately, below, and then present a conceptual model for how these factors interrelate to create therapeutic itineraries in southwestern Uganda.

##### A. Traditional healers care about their patients

Patients recruited from traditional healers report positive experiences with their care, specifically describing that treatments effectively relieve their symptoms. Participants state that they prefer traditional therapies because traditional practitioners “heal faster”. This efficient healing is sometimes attributed to the fact that traditional practitioners spend more time personally treating and caring for their patients, compared with healthcare workers in biomedical settings:

> *Those* [*bonesetters*] *are super! They heal faster than biomedicals. When you take your patient to a bonesetter, he does not take long to get healed, compared to one in the hospital. In hospitals, the healing process is long because they do not do much more than hanging you there* [*in traction*] *and leave you. You can even become lame because they do not check to see whether you are healing or not. But for the healer, he does his reviews* [*checks your wound healing*] *constantly*. (*Traditional healer patient, female, 68 years old*)

Patients receiving traditional care also state that they are treated with respect when visiting healers, and that healers are motivated to care for patients, rather than being strictly economically driven. Participants reported that healers attend to patients immediately, even if they did not have money; a few participants stated that healers allowed them to pay for services rendered in installments, or in kind (through farm goods). A participant seeking care from a traditional birth attendant described her preference for traditional healing, emphasizing the kindness of her practitioner:

[*The healer*] *does everything for you. Her services are excellent. In fact, when you deliver* [*your children*] *from here, you do not even think of going elsewhere another time. She cares so much about her clients. In fact, for all my pregnancies, I received antenatal care from this healer. She is my neighbor, and instead of going to sit at the hospital the whole day waiting for checkup, I come here. She is my neighbor and her services are good. So, I come get my antenatal checkup, and go back home to do my chores*. (*Traditional healer patient, female 35 years old*)

In contrast, patients describe experiences with biomedicine with narratives of disrespect, mistreatment, neglect or “abuse”. The central message of these biomedical testimonies is that healthcare workers do not care about their patients. In some cases, participants referred to these accounts while explaining why they tend to avoid biomedical facilities. A woman describes her experience receiving antenatal care at the local hospital:

> *I came to this hospital for antenatal care and found a nurse who treated me badly. She would tell you to lay on the bed and instead of telling you what to do, she would shout at you and say, “Don’t face me! Face the other side!” in a loud voice, and you wonder what the problem was. She embarrassed me and I felt ashamed. I promised myself never to return in this hospital …. She would only shout at us. She was horrible*. (*Biomedical patient, female, 38 years old*)

A number of participants describe experiences at biomedical facilities where they are never attended to by biomedical staff, despite waiting for many hours – sometimes spending the entire day without receiving medical attention. These hours spent waiting come at the expense of childcare, household duties and income-generating activities. One man describes his experience seeking biomedical care for a toothache as follows:

> *I went to the referral hospital and spent there the whole day without treatment. The following morning, when I went back, I was given only Panadol* [*Acetaminophen*]. *I felt so sad*. (*Biomedical patient, male, 56 years old*)

Another patient states that he gave up after waiting all day for a voluntary circumcision procedure:

> *You reach there and sit for the whole day without treatment. Drugs are never there and health workers do not attend to patients as it should be. They arrive at work late and leave work early. They are really bad. I went* [*to the clinic*] *one time for circumcision and sat there for many hours until I got hungry and gave up. I left without seeing any doctor*. (*Traditional healer patient, male, 27 years old*)

##### B. Biomedicine uses modern technologies to heal

Participants state that biomedical care is preferred in instances where “modern” technologies can be utilized to provide a diagnosis for one’s symptoms, and guide treatment. Through blood and radiological tests, healthcare providers can identify the specific cause of a patient’s illness, and provide appropriate care. Patients perceive that the information generated by biomedical technology validates the therapies administered to them:

> *They use machines to diagnose and test for conditions. The give the right medical information*. (*Biomedical patient, male, 25*).

Having received a specific diagnosis, participants also believe that the treatment recommended by healthcare workers will be effective in alleviating their symptoms. For example, one participant described how appropriate medicines have the capacity to heal, even if taken in small amounts:

> *When you come* [*to the clinic*] *you get diagnosed and they write for you a prescription and you get the medicine then their service is good … Even if you get very little medicine from them and take it, you get healed*. (*Biomedical patient, female, 60 years old*)

Another patient explains why the capacity to intervene with modern biomedical technology is more effective in treating symptoms than traditional medicine:

> *Biomedical facilities are good … when you are, for instance, in a critical condition, they can put you on life support machines, or they can put you on a drip. They can also give you tablets and injections that can help you. Traditional healers can’t manage something like that. They don’t have modern equipment. They don’t have tablets, and they don’t’ have drips and injections*. (*Traditional healer patient, male, 26 years old*)

Results from biomedical testing guide what some participants describe as “proper”, effective treatment, compared with traditional healing where therapies are provided in the absence of any diagnostic testing:

> [*Biomedical facilities*] *diagnose you and inform you of the ailment that you are suffering from, and at times inform you that your health is okay … When you visit biomedical health facilities they diagnose you and inform you of your results and in case you are HIV positive, you can start on medicine …* [*Traditional healers*] *don’t have equipment to diagnose, so how do they diagnose for conditions? … I don’t trust them*. (*Biomedical patient, Female, 22 years old*)

While biomedicine is favored for its use of diagnostic technologies, other participants describe preference for traditional healing *specifically because* these approaches could enable avoidance of biomedical procedures, which participants describe as “unnecessary” and having high morbidity and mortality. Participants state that an advantage of traditional healing is that it supports the body to heal “naturally”, rather requiring modern, invasive interventions. Participants report seeking traditional care after having been told by biomedical providers that they would require an operation in order to recover. Those who ultimately healed after receiving traditional care declared that biomedical providers rush to use modern technologies, instead of allowing the body to heal on its own. One patient describes his experience receiving care from a bonesetter, after suffering severe extremity fractures after falling from a motorcycle:

> [*The hospital staff*] *told me that the doctors will cut off my leg because it was badly injured and that there was no way they could fix it … When we reached* [*this healer*], *they told me that the bone that joins the knee was broken but promised that since I was in that place, in two to three weeks, I will be able to walk again. They then aligned my leg and started the treatment … I am now getting better. If I had remained at the hospital, I know my leg would have been cut off by now*. (*Traditional healer patient, male, 35 years old*)

Another patient describes how effective treatment from an herbalist allowed her sister to avoid a Caesarean section with her twin pregnancy:

> *These healers are very useful … my elder sister had a problem with her twin pregnancy. She was stuck with the pregnancy because the babies could not move. They took her to one of the traditional healers and was given medicine which helped her so much and she delivered her babies without difficulties. We thought she would be operated on while giving birth* [*via Caesarean section*] *because the doctors at referral hospital had told her that she will not manage to push and advised her to go for an operation, which did not happen because of the medicine the healer gave her*. (*Traditional healer patient, female, 30 years old*)

Participants described fear of utilizing biomedical facilities to deliver their children, as they believed that physicians would perform unnecessary Caesarian sections, considered a high-risk procedure for both mothers and infants:

[*Doctors*] *rush women to the operating theatre when it’s not necessary. Many women and babies have lost their lives due to the negligence of doctors. Women fear to deliver from hospital*. (*Traditional healer patient, male, 26 years old*)

##### C. Peer “testimony” influences healthcare engagement

Our participants recount social narratives, or “testimonies” which describe healthcare experiences among peers within their communities. These discursive events evaluate a provider’s competence and effectiveness in addressing ailments, and describe negative or positive outcomes of treatments. Participants indicate that peer testimonies strongly influence where they choose to seek care for their symptoms. We found that biomedical narratives frequently reinforced individual reports of mistreatment; in contrast, narratives about traditional healing were generally positive and affirmed the “real” nature of this form of healthcare.

Numerous participants who received care from traditional healers describe negative peer narratives about biomedicine. A participant describes the testimony from his neighbor that influenced his decision to seek care from a traditional bonesetter:

> *My neighbor reached* [*the referral hospital after injuring his leg*], *but nothing much was done. They made him sit on the waiting bench and the doctor told the caretaker to go and buy a bandage and find an empty box. The doctor then dismantled the box and tied it on the leg using the bandage and left him there. He remained there until morning. …. He never got any treatment* [*for the leg injury*] *apart from the empty boxes they tied on the leg. I will never forget what he experienced from the referral hospital. It was so bad and so discouraging. Health workers do not care about patients*. (*Traditional healer patient, male, 57 years old*)

A number of participants recalled community narratives indicating that healthcare workers would intentionally withhold treatment or harm their patients. One woman seeking care at a traditional birth attendant practice describes stories that made her fear that she would be harmed at the hands of healthcare workers:

> *There was a woman in labor who was supposed to be taken to the operating theatre but the nurses asked her for money, which she did not have. They refused to work on her until other patients contributed some money and gave it to the nurses … Those nurses do not mind whether you die from there or not … There is also one mother I heard about who took her child for immunization and got an argument with the nurse. Intentionally the nurse gave the child overdose and the child died. Some of these health workers are so wicked*. (*Traditional healer patient, female, 35 years old*)

Negative peer testimonies were not limited to patients of healers. For example, one woman seeking biomedical care told a story about her neighbor suffering mistreatment at the same facility.

> *My pregnant neighbor delivered her baby in the village compound*. [*When they arrived at this hospital for post-partum care*], *the nurse abused her, saying that she should take her stupidity back to her village. They do not care*. (*Biomedical patient, female, 22 years old*).

In stark contrast to narratives surrounding biomedical care, peer testimony surrounding traditional healing is largely positive. Healers are lauded for their effective care, and patients are guided by peer testimonials in selecting which healer to visit for their ailments. One participant seeking care at a traditional herbalist describes the impact of peer endorsements on her decision to seek care from this particular healer:

> *This healer is popular and well known, and wherever you go, people will recommend her to treat your sick child … I have seen so many different people come here to receive treatment … I am impressed*. (*Traditional healer patient, male, 18 years old*).

A central concept in many testimonies about traditional medicine is the genuineness of the healer, and how they should be set apart from traditional healers who may be “fake” or “quacks”. One participant describes how testimonies from peers with similar injuries directed him to seek care from a specific bonesetter, and how testimonies generate more patients for particular healers:

> *Most traditional healers are quacks, and personally I don’t trust them*. [*Interviewer: Then how do you know that you will heal from this treatment?*] *I get the confidence from other people who have been treated here. There is a man from a nearby dairy. He bones were more severely broken than mine, but he healed from here, and is now doing his work. I have heard many people’s testimonies that they have been healed from here … When I come here and get healed, I will direct another one because he will be healed too and that person will also direct others… A healer who is real does not need to advertise on the radios because the people they heal create market for them*. (*Traditional healer patient, Male, 26 years old*)

## DISCUSSION

This study identified factors that drive engagement with healthcare resources in a medically pluralistic setting, and identified three central factors that contribute to therapeutic pluralism. These factors may be summarized as follows: 1) traditional healers care about their patients, while biomedical providers do not; 2) biomedical technologies can provide diagnosis and guide treatment, but these technologies are sometimes intentionally avoided; and 3) peer testimonies influence healthcare utilization, largely in favor of traditional healing. Figure 1 presents a conceptual model integrating our findings to show how influences at the <u>healthcare provider</u>, <u>healthcare system</u>, and <u>peer</u> levels influence individual engagement in pluralistic settings. This model is not inclusive of all variables that influence health engagement, but illustrates categories that were described by our participants in driving their own healthcare decision making, specifically regarding decisions to utilize traditional or biomedical care.

First, our data illustrate that <u>healthcare provider characteristics</u> are of central importance to patients. Specifically, the quality of interpersonal interactions can either motivate or deter engagement with healthcare services. In our study, patient-provider interactions with traditional healers are described as generally respectful and supportive. In contrast, patient-provider interactions in biomedical contexts included narratives of neglect and “abuse”. The health effects of negative interactions with biomedical staff have been well described in cases of disengagement with HIV care among people living with HIV[33- 35], decreased PrEP utilization among key populations[36] and among women giving birth[37-39]. Other researchers have similarly shown that traditional healers are favored in some cases because they provide social support within their communities, functioning as counselors, social workers, spiritual guides, and legal advisors[5,20,25,40-44].

Characteristics of the available <u>healthcare systems</u> impact healthcare engagement. Participants appreciate that biomedical laboratory and radiologic testing guide diagnosis and treatment, thereby gaining reassurance that they can heal from their illness through “proper” treatment. We note that the desire for healthcare directed by test results is the central factor favoring biomedical healthcare utilization among our participants.

Interestingly, data from high-resource contexts has shown that diagnostic test results do not increase patient reassurance or decrease health-related anxiety in outpatient biomedical settings[45,46]. It is likely that in our medically pluralistic study site, the capacity of biomedical facilities to perform diagnostic testing is distinctive in contrast to traditional healing approaches, and therefore considered a benefit. Further, our qualitative data draws from patients’ own words describing reassurance in receipt of diagnostic testing, whereby the prior studies employ quantitative measurements of patient reassurance and anxiety.

We also found that traditional healthcare is sometimes preferred as a means to avoid invasive procedures, such as orthopedic fixation, limb amputation, or Caesarean section. Our findings are congruent with prior research demonstrating avoidance of facility-based obstetric services, preference for traditional home birth[25,39,47], and bonesetters to heal orthopedic injuries in sub-Saharan Africa[48,49]. Motivation to avoid invasive operative procedures is further explained by data that show poor post-operative outcomes throughout sub-Saharan Africa[50]; for example, maternal mortality after Caesarean section is fifty times higher in Africa compared with high income countries[51]. As such, patients consider invasive biomedical procedures high risk, and seek to avoid them through receipt of traditional therapies.

Additionally, the content of <u>peer</u> testimonies strongly influences patients’ decisions to utilize traditional or biomedical care. Peers can be defined as other adults residing in the same community as the participant, who have relevant experiences receiving biomedical care, traditional care, or both. Peer influences have been shown to have strong impact on individual healthcare engagement in the cases of HIV services utilization[52-54], adolescent health[55,56], mental health[57], and substance misuse[58], for example.

Our study shows how peer testimonies serve as endorsements of traditional healing, legitimizing its use through descriptions of clinical effectiveness. In contrast, largely negative narratives regarding biomedicine potentiate avoidance of these facilities and services.

Finally, our data contribute to a growing body of work that emphasizes the important role of traditional healers within the communities they serve. Our findings illustrate why traditional medicine may be preferred, even when biomedical services are available and accessible to patients. Lack of biomedical engagement in pluralistic settings should not simply be attributed to lack of access, but should be considered an individual’s informed healthcare choice. We suggest that public health interventions specifically engage with traditional healers to increase intervention impact and community acceptability; they are well positioned allies for any community-based health program. Studies have shown that healers are interested in working with biomedical providers to improve health outcomes for their patients[59-61].

There are a few limitations of this study. It is beyond the scope of this research to investigate the effectiveness or appropriateness of therapies administered by providers to the participants in our study. Similarly, it is out of our scope to consider the ethnopharmacological and ethnobotanical literature investigating the clinical efficacy of traditional therapies, which could impact a patients’ clinical improvement and assessment of effective treatment; that literature is not discussed here. Last, qualitative data are meant to be specific and contextual rather than broadly generalizable, and are useful in generating hypotheses. As such, our data suggest numerous directions for future study; for example, do the factors we identified influence patients differentially as a function of patient age, gender, socio-economic status, or other individual characteristics? Are there distinctions among healers or their clients that predict increased biomedical or traditional utilization, such as gender, specialty, symptoms, or cost? How can public health initiatives that collaborate with traditional healers be optimally delivered in medically pluralistic settings?

## CONCLUSIONS

Patients perceive clear advantages and disadvantages to biomedical and traditional care in medically pluralistic settings. We identified factors at the healthcare provider, healthcare system, and peer levels which can influence patients’ therapeutic itineraries, and illustrate why traditional healers are sometimes preferred. Our findings provide a basis for public health interventions in medically pluralistic communities, and underscore the importance of recognizing and engaging with traditional healers as important stakeholders in community health.

## Data Availability

Deidentified data may be shared upon reasonable request by emailing the first author.

## ACKNOWLEDGEMENTS

The authors would like to acknowledge the work of our research assistants, Patricia Tushemereirwe, Donny Ndazima and Doreen Nabukalu. We are grateful to the traditional healers and patients in Mbarara District for sharing their experiences with us.

## CONTRIBUTORSHIP STATEMENT

RS conceived of the study. RK and JMA provided input on study design, study procedures. RS and JMA oversaw data collection. RS was primarily responsible for data analysis, with input from JMA, RK and NW. RS composed the first draft of the manuscript. All authors provided input and approve of the final submission.

## DATA SHARING STATEMENT

Deidentified data may be shared upon reasonable request by emailing the first author.

## PATIENT AND PUBLIC INVOLVEMENT STATEMENT

Patients were included as participants in this study. They did not directly participate in the design or implementation of the study, as the purpose of the study was to elicit patient perspectives on community healthcare resources. Results of this study were used to guide development of a study community advisory board, which includes patients and other stakeholders, including healthcare providers and community leaders.

